# COVID-19 onset reduced the sex ratio at birth in South Africa

**DOI:** 10.1101/2022.02.07.22270630

**Authors:** Gwinyai Masukume, Margaret Ryan, Rumbidzai Masukume, Dorota Zammit, Victor Grech, Witness Mapanga

## Abstract

**Background:** The sex ratio at birth [defined as male/(male+female) live births] is anticipated to approximate 0.510 with a slight male excess. Following sudden unexpected stressful events, this ratio has been observed to decrease transiently around 3-5 months following such events. We hypothesised that stress engendered by the onset of the COVID-19 pandemic may have caused such a decrease in South Africa 3-5 months after March 2020 since in this month, South Africa reported its first COVID-19 case, death and nationwide lockdown restrictions were instituted.

**Methods:** We used publicly available recorded monthly live birth data from Statistics South Africa. The most recent month for which data was available publicly was December 2020. We analysed live births for a 100-month period from September 2012 to December 2020, taking seasonality into account. Chi-squared tests were applied.

**Results:** Over this 100-month period, there were 8,151,364 live births. The lowest recorded monthly sex ratio at birth of 0.499 was in June 2020, 3 months after March 2020. This June was the only month during this period where the sex ratio inverted i.e., fewer male live births occurred. The predicted June 2020 ratio was 0.504. The observed June 2020 decrease was statistically significant p = 0.045.

**Conclusions:** The sex ratio at birth decreased and inverted in South Africa in June 2020, for the first time, during the most recent 100-month period. This decline occurred 3 months after the March 2020 onset of COVID-19 in South Africa. As June 2020 is within the critical window when population stressors are known to impact the sex ratio at birth, these findings suggest that the onset of the COVID-19 pandemic engendered population stress with notable effects on pregnancy and public health in South Africa. These findings have implications for future pandemic preparedness and social policy.

## Introduction

The sex ratio at birth (SRB) also known as the secondary sex ratio is defined as [male/(male+female) live births] [1]. At population level it approximates 0.510 with male live births slightly exceeding female live births. The SRB may act as a sentinel health indicator underscoring adverse circumstances through its decrease, but also highlighting improved circumstances with its increase [2, 3]. Following sudden unexpected stressful events, the population SRB has been observed to decrease transiently usually some 3-5 months following events such as the death of a highly popular public figure [4], terrorist attacks [5] and wide spread protesting/rioting [6]. Such stressful life events impact on maternal neuroimmunoendocrine condition; male fetuses are more vulnerable to such changes in condition [7]. Another potential window is 9 months after such events as was the case in Japan when the SRB dipped 9 months after the Kobe earthquake [8]. However, these windows of SRB decline are not a universal phenomenon due to varying contextual factors [9, 10].

It has been proposed that COVID-19 stress might result in SRB decline [11]. In March 2020, South Africa reported its first known polymerase chain reaction (PCR) confirmed COVID-19 case [12]. During that same month the World Health Organization declared COVID-19 to be a pandemic [13], and this was accompanied by a surge in local and global media attention on COVID-19 [14]. In addition, during March 2020, South Africa declared a national state of disaster and instituted nationwide lockdown measures which included school closures, and restrictions on gatherings, domestic and international travel [15]. Furthermore, the first death attributed to COVID-19, in South Africa also occurred [16]. In short, March 2020 was a momentous month in relation to COVID-19 in South Africa.

Our primary hypothesis was that stress engendered by the onset of the COVID-19 pandemic might have caused a transient decrease in SRB in South Africa 3-5 months after March 2020. The secondary hypothesis was that an SRB dip occurred 9 months later, in December 2020.

## Materials and Methods

### Data and statistical analysis

Monthly male and female recorded live births, from the national birth registration system, were obtained from Statistics South Africa’s publicly available annual reports [17]. For the intercensal period 2011–2016, completeness of birth registration was estimated at 88.6%. Data was extracted for a 100-month period from September 2012 to December 2020. The last year the SRB, in South Africa, was known to have been influenced by an event was in 2011 [18]. December 2020 was the most recent month for which data was available. We conducted post-hoc analysis considering 3-5 and 9 months after the April 2015 protests/riots [19], as the SRB might have been affected consequent to stress induced by this event.

To take seasonality into account, similar months across the years were compared. Restriction [20] was used to control for potential confounding factors. Chi-squared tests were used. This methodology is an alternative to time series analysis that has previously been used with similar South African data [21]. P < 0.05 was considered to be statistically significant. For analysis, Stata version 17BE (College Station, TX) was used.

### Ethical considerations

Due to the anonymized nature of the data, ethical approval was not required.

## Results

Over this 100-month period, from September 2012 to December 2020, there were a total of 8,151,364 live births as 4,113,288 males and 4,038,076 females, SRB 0.505. The lowest recorded monthly sex ratio at birth, during the 100-month period, was in June 2020 at 0.499 (Fig 1). June 2020 was also the only month during this most recent 100-month span that the SRB inverted i.e., fewer male than female live births occurred. The SRB for December 2020 was 0.503. Based on similar months in previous years, the moving average approach predicted an SRB of 0.504 for June 2020 (Fig 2) and 0.505 for December 2020 respectively. The June 2020 SRB decrease was statistically significant p = 0.045, while for December 2020 there was no statistically significant change p = 0.293 (Table 1). Post-hoc analysis, S1 Table, revealed no statistically significant result.

**Fig 1.**
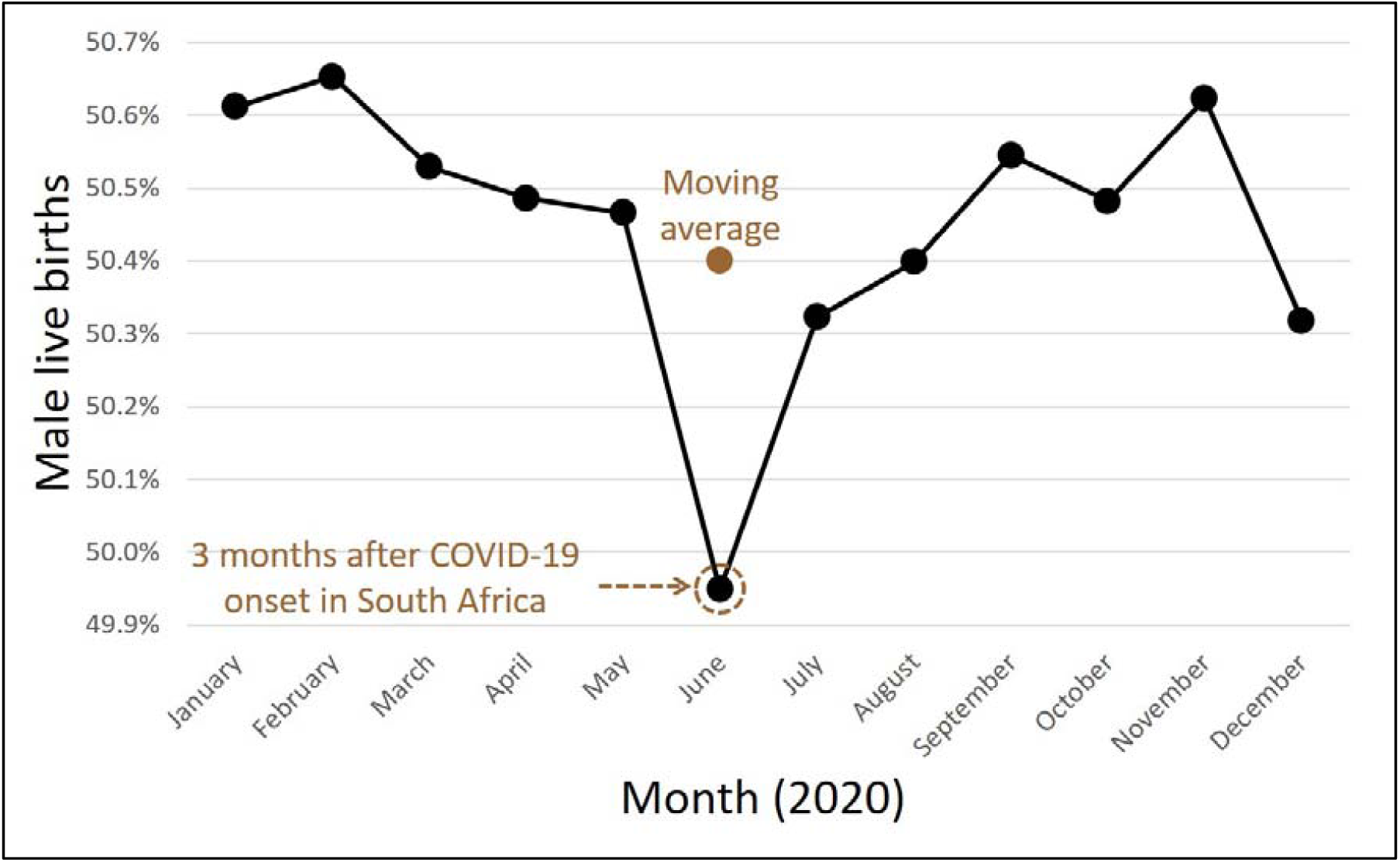
Monthly proportion of male live births in 2020.

**Fig 2.**
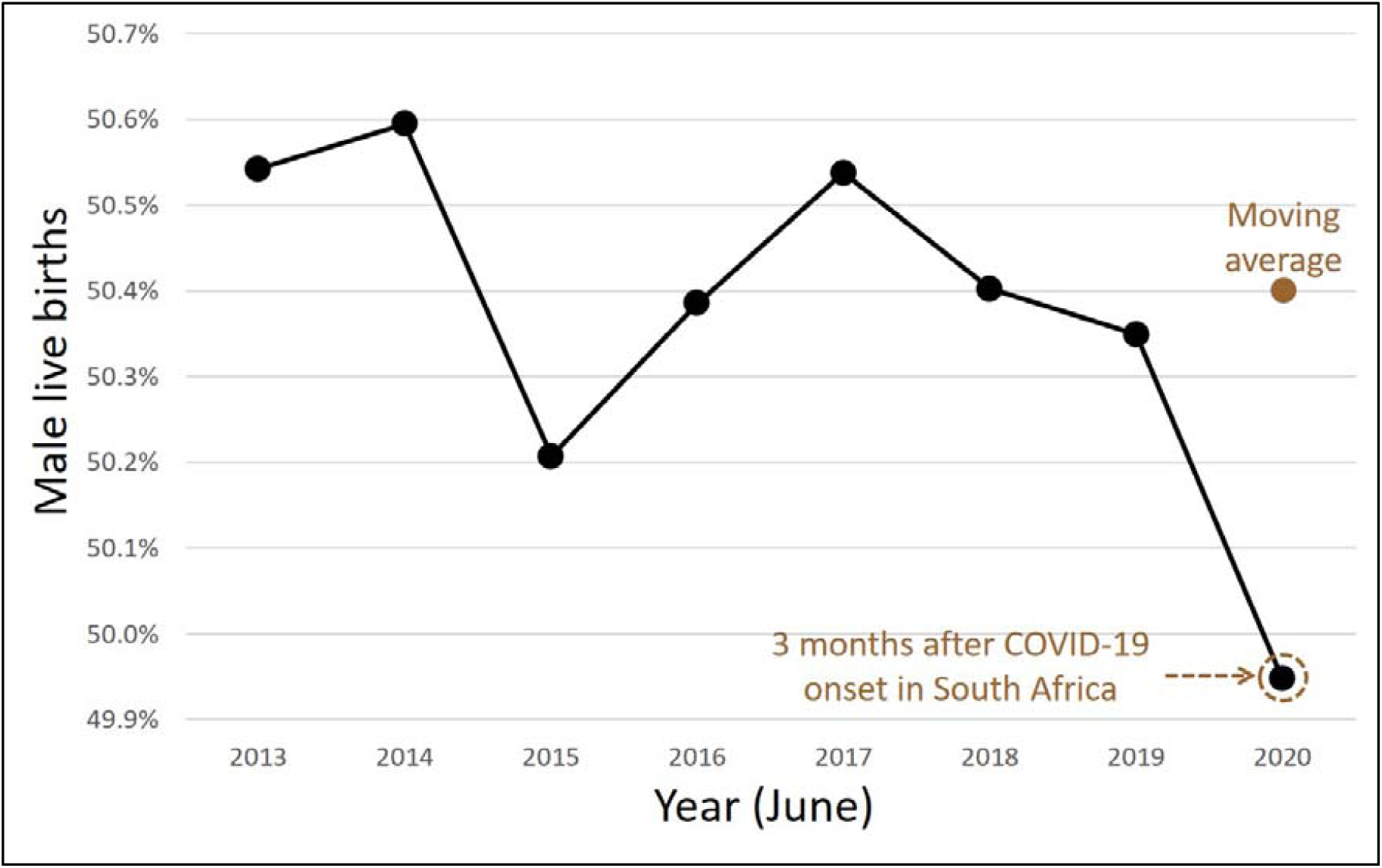
Proportion of male live births during the month of June from 2013-2020.

**Table 1.** Sex ratio at birth restricted from 2018-2020.

| Period | Male live births | Female live births | Sex ratio at birth |
| --- | --- | --- | --- |
| June 2018 + 2019 | 81,078 | 79,870 | 0.504 |
| June 2020 | 42,022 | 42,109 | 0.499 |
|  | Pearsons's Chi-squared = 4.0292 |  |  |
|  | P = 0.045 |  |  |
| December 2018 + 2019 | 81,237 | 79,495 | 0.505 |
| December 2020 | 41,795 | 41,268 | 0.503 |
|  | Pearsons's Chi-squared = 1.1058 |  |  |
|  | P = 0.293 |  |  |

## Discussion

In this study, we provide insight into how the SRB, in South Africa, was influenced by the onset of the COVID-19 pandemic. Over the study period of 100 months, we observed the lowest SRB of 0.499 in June 2020, 3 months after the first COVID-19 case, death and lockdown occurred in March 2020. Notably, the SRB had inverted i.e. fewer male live births occurred. This is the only month this happened during the 100-month period, from September 2012 through to December 2020. Our findings are inline with the hypothesis that COVID-19 stress would result in SRB decline [11], driven by the Trivers-Willard hypothesis which posits that adverse environmental conditions can lower the ratio of males to females [22]. Our findings are also consistent with studies that have reported a transient SRB decline 3-5 months following unexpected events which stressed populations acutely and severely [4-6]. Although an SRB decline has been reported 9 months after an acute unanticipated severe stressor [8], we did not find evidence of this in South Africa, and the SRB, in December 2020, was not appreciably different in comparison to previous Decembers.

The timing of the SRB dip, 3 months following the onset of COVID-19 in South Africa suggests a mechanism operating in ongoing pregnancies whereby, due to stress, there is an elevated risk of fetal death among males [23] resulting in fewer live born males. Thus beyond the direct adverse effects of the pandemic pathogen on pregnant women evidenced by a higher maternal mortality rate among women admitted with SARS-CoV-2 infection [24], anxiety and stress likely generated by news of the impending pandemic [14], in March 2020, led to disproportionate male fetal loss in June 2020. Triangulation with antepartum deaths from South Africa’s perinatal mortality audit system, when the data becomes publicly available, would be needed to directly confirm this excess male fetal loss [25]. Had a dip been witnessed in December 2020, 9 months after the onset of COVID-19, this would have pointed to a conception mechanism.

It is plausible that the June 2020 SRB decline could have been attributed to an event approximately 9 months earlier [8], in September 2019. Indeed there were protests/riots during that September which garnered significant media attention [26]. However, several factors are not consistent with this paradigm of the South African June 2020 SRB decrease being primarily related to September 2019 events. First, there is the precedent of the Los Angeles protests/riots [6] where the SRB perturbation occurred during the 3-5 month post-event window. Second, similar protests/riots to the aforementioned ones have occurred before in South Africa during the most recent 100-month period, in April 2015 [19], but with no significant SRB decline either 3-5 or 9 months later (post-hoc analysis). Third, contrary to COVID-19, which had broader nationwide ramifications, these protests [6, 19, 27] were confined to certain locations with possibly more localised effects.

Ecological fallacy, the limited ability to extrapolate findings to the individual level applies to this study [28]. Nonetheless, the ecologic study technique is arguably best suited to investigate how a population outcome is influenced by a population stressor [29]. In late March 2020, due to the institution of nationwide lockdown measures, the Department of Home Affairs (DHA) only offered limited civic services. Due to this, there was a decline in the number of live births registered early (< 30 days). With the easing of restrictions, services resumed in early May 2020. In 2020, there was an overall compensatory increase in the number of live births registered late (> 30 days) [17]. The neonatal mortality rate, death within the first 28 completed days of life, in South Africa approximates 12 deaths per 1,000 live births [30]. Nonethless, the total recorded live births for 2020 is inline with prior years. Overall, this means when restrictive measures were lifted those who had missed registering a live birth came forward. Thus the present analysis is unlikely to have been affected by closure of the DHA offices necessitated by lockdown regulations.

The present analysis covers the first COVID-19 wave in South Africa. At the time of writing, South Africa had experienced four waves [31]. It is possible that the SRB may have been transiently perturbed in the following waves. This is an area for research when the recorded live birth data for these periods becomes available.

## Conclusions

The sex ratio at birth decreased and inverted in South Africa in June 2020, for the first time, during the most recent 100-month period. This decline occurred 3 months after the March 2020 onset of COVID-19 in South Africa. As June 2020 is within the critical window when population stressors are known to impact the sex ratio at birth, via elevated risk of fetal death among males, these findings suggest that the onset of the COVID-19 pandemic engendered population stress with notable effects on pregnancy and public health in South Africa. Our findings can help inform future pandemic preparedness [32] and social policy [33]. Whether other countries experienced a decline in the SRB, following the onset of COVID-19 is an area for future research.

## Data Availability

All data produced in the present work are contained in the manuscript

http://www.statssa.gov.za/publications/P0305/P03052020.pdf

## Acknowledgements

We acknowledge Statistics South Africa for the recorded live birth data.

## Author Contributions

Conceptualization: GM, MR, RM, DZ, VG, WM

Data curation: MR, GM

Formal analysis: GM, DZ

Writing – original draft: GM

Writing – review & editing: MR, RM, GM, DZ, VG, WM

---

**S1 Table.** Sex ratio at birth restricted from 2013-2016.

| **Period** | **Male live births** | **Female live births** | **Sex ratio at birth** |
| --- | --- | --- | --- |
| August 2013 + 2014 | 87,302 | 85,901 | 0.504 |
| August 2015 | 40,335 | 39,777 | 0.503 |
|  | Pearsons's Chi-squared = 0.0691<br>P = 0.793 |  |  |
| January 2014 + 2015 | 90,339 | 88,510 | 0.505 |
| January 2016 | 39,756 | 38,441 | 0.508 |
|  | Pearsons's Chi-squared = 2.3632<br>P = 0.124 |  |  |

